# Development and Validation of the Alimetry® Gut-Brain Wellbeing Survey: A novel patient-reported mental health scale for patients with chronic gastroduodenal symptoms

**DOI:** 10.1101/2023.09.25.23296105

**Authors:** Mikaela Law, Isabella Pickering, Gayl Humphrey, Gabrielle Sebaratnam, Gabriel Schamberg, Katie Simpson, Chris Varghese, Peng Du, Charlotte Daker, I-Hsuan Huang, Sahib S. Khalsa, Armen Gharibans, Greg O’Grady, Christopher N. Andrews, Stefan Calder

**Author notes:** **Corresponding Author:** Dr Stefan Calder, Department of Surgery, The University of Auckland, Auckland, New Zealand.

## Abstract

**Objective:** There is currently a lack of validated questionnaires designed specifically to assess mental health within patients with chronic gastroduodenal symptoms. This research describes the multi-phase process used to develop and validate a novel mental health scale for patients with chronic gastroduodenal symptoms, the Alimetry® Gut-Brain Wellbeing (AGBW) Survey.

**Methods:** A patient-centred multi-phase process was implemented. In Phase 1, the most relevant concepts for this patient population were selected from existing mental health scales, using data from 79 patients. In Phase 2, an interdisciplinary panel of experts generated scale items. In Phase 3, the scale underwent pre-testing with gastroenterologists (n=9), health psychologists (n=3), and patients (n=12), with feedback incorporated over multiple rounds. Lastly, the psychometric properties of the scale were assessed in a sample of 311 patients via an online survey.

**Results:** The AGBW Survey comprises a patient preface, 10 close-ended questions, and an optional open-ended question. This multidimensional scale assesses general mental health, alongside specific subscales relating to depression, stress, and anxiety. The subscale and total scores demonstrated high internal consistency (α= .91 for the total scale; α= .72-.86 for subscales) and good convergent, divergent, concurrent validity, and known groups validity, with large effect sizes.

**Conclusions:** The AGBW Survey is a brief, valid, and reliable scale for assessing mental health in patients with chronic gastroduodenal symptoms. It can be used as a tool to complement physiological tests and has the potential to guide psychological referrals, inform multidisciplinary management, and evaluate treatment outcomes.

## Introduction

Chronic gastroduodenal symptoms, like those experienced in gastroduodenal disorders of gut-brain interaction (DGBIs), affect more than 10% of the global adult population and constitute a significant health issue due to their increasing prevalence and rising healthcare costs[1–4]. These symptoms include chronic nausea, vomiting, belching, regurgitation, epigastric pain/burning, early satiation, and excessive fullness, which are often experienced in conditions such as functional dyspepsia, gastroparesis, and chronic nausea and vomiting syndrome[2–4]. The management and diagnosis of these symptoms present considerable challenges as such patients frequently exhibit overlapping symptomology devoid of identifiable structural aetiology[5,6]. Consequently, there are limited well-defined diagnostic pathways and targeted treatment approaches[4,7,8].

Growing evidence shows a bidirectional relationship between gastrointestinal symptoms and psychological factors. Psychological comorbidities are common in patients with chronic gastroduodenal symptoms, and stress, anxiety, and depression have been found to trigger and exacerbate gastrointestinal symptoms[8–14]. The gut-brain axis, a complex neurohormonal pathway that facilitates communication between the brain and the gastrointestinal tract, plays a crucial role in this association[9,15–17]. Psychological interventions have the potential to improve mental wellbeing and gastrointestinal symptoms in these patients[14,18–21]. Therefore, early identification and management of psychological comorbidities are crucial for significant improvements in gastrointestinal symptoms and quality of life.

The growing recognition of the gut-brain axis and the adoption of a biopsychosocial framework have led to the recommendation of psychological assessments as a critical part of the standard care for managing chronic gastroduodenal symptoms[3,9,13,22]. Clinicians routinely ask mental wellbeing questions when evaluating gastrointestinal patients; however, these questions are often informal[23–25]. Although validated mental health questionnaires exist, these have limitations when used within this patient population. For example, many questionnaires assessing depression and anxiety include questions on physical symptomatology, such as reduced appetite and disrupted sleep. However, in patients with chronic gastroduodenal symptoms, these physical manifestations may be primarily related to their gastrointestinal disorder. Therefore, these questions may not be a valid reflection of their mental health, potentially resulting in an overestimation of psychological concerns and inaccurate formulations of their symptomatology, which may lead to ineffective management[13].

To date, no general mental health scale has been developed and validated among patients with chronic gastroduodenal symptoms. The ongoing improvement and validation of psychometrics within novel samples is recommended to increase the dependability and validity of the results[26]. Therefore, there is a need to develop a brief self-report scale specifically designed for use in patients with chronic gastroduodenal symptoms to ensure valid and reliable assessments of mental health. Here we describe the steps used to develop and validate a novel mental health scale, the Alimetry® Gut-Brain Wellbeing (AGBW) Survey for patients with chronic gastroduodenal symptoms.

## Methods

As shown in Figure 1, the AGBW Survey was developed and validated in four mixed-methods phases, with guidance from the results from a precursory user needs interview study with patients with gastroduodenal DGBIs and gastroenterology clinicians[25]. Each phase involved co-design with gastroenterologists, psychogastroenterologists, and patients with chronic gastroduodenal symptoms to ensure face and content validity, comprehensibility, and acceptability.

**Figure 1.**
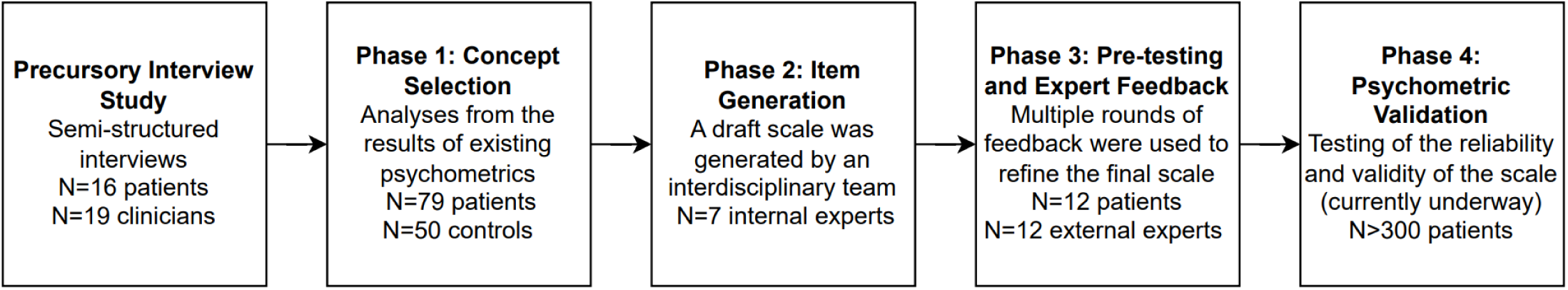
Flowchart of the phases used to develop the Alimetry Gut Brain Wellbeing Survey.

### Phase 1. Concept Selection

The first phase aimed to identify the most important mental health concepts to include in the new scale. Valid concepts were selected from existing mental health scales, using expert feedback and the analysis of psychometric data from a sample of patients with chronic gastroduodenal symptoms, to reduce the number of constructs to those most relevant and contextualized to the target patient population. The full methods for concept selection are provided in the Supplementary Methods.

### Phase 2. Item Generation

An interdisciplinary panel of experts (including two health psychology researchers specialising in psychogastroenterology, a gastroenterologist, a gastrointestinal surgeon, a digital health translational researcher, and two bioengineers specialising in gastric electrophysiology) generated novel draft items based on the concepts identified from Phase 1.

### Phase 3. Pre-Testing and Expert Feedback

The draft scale was then pre-tested with a sample of patients with chronic gastroduodenal symptoms and reviewed by independent external experts to ensure the acceptability, clarity, comprehensibility, and content and face validity of the scale.

### Sample

The pretesting sample consisted of 12 patients with chronic gastroduodenal symptoms (11 females; mean age= 33.3 years, age range= 20-55 years). All patients met the Rome IV criteria[27] for functional dyspepsia, with 10 also having a coexisting diagnosis of chronic nausea and vomiting syndrome and nine also having a diagnosis of gastroparesis. The external experts comprised key opinion leaders, including nine gastroenterologists, one health psychologist specialising in psychogastroenterology, and two health psychology researchers. Patients and experts were recruited until data saturation.

### Procedure

Experts and patients viewed the draft scale alongside images that showed potential implementation on a tablet interface. They answered a series of open-ended questions, including about the acceptability of the scale items, the utility of the scale in clinical practice, the ease of understanding and comprehension of the question wording, and whether the scale could be improved. This feedback was incorporated into the scale and sent back to respondents for further feedback. This process occurred until all reviewers were satisfied with the scale wording.

### Phase 4. Psychometric Validation

Psychometric validation of the scale was conducted using an anonymous, cross-sectional survey of patients with chronic gastroduodenal symptoms. All patients provided informed consent. Ethical approval was granted by the Auckland Health Research Ethics Committee Application (AH25798), and the trial was pre-registered at ANZCTR.org.au (ACTRN12623000385640).

### Sample

Patients were recruited via convenience sampling through social media, clinic flyers, and clinic lists. Patients were included if they were over 18 years old and able to speak, read, and write fluently in English. Patients also had to meet the Rome IV criteria[27] and/or have a self-reported clinical diagnosis for at least one of the following conditions: gastroparesis, functional dyspepsia, chronic nausea and vomiting syndrome, cyclic vomiting syndrome, rumination syndrome, cannabinoid hyperemesis syndrome, or a belching disorder. Clinician confirmation of diagnosis was not collected. Vulnerable participants and patients with self-induced vomiting or an eating disorder were excluded. Patients were recruited globally and efforts were made to ensure adequate recruitment across geographic regions, conditions, and genders.

### Procedure

The anonymous survey was completed online via Qualtrics (Qualtrics, Provo, UT) and took approximately 15 minutes. Patients were presented with a demographics questionnaire and a battery of psychological questionnaires, including the AGBW Survey, presented in a randomized order. The survey ended with an optional feedback form about the scale and participants were provided with the option to enter a prize draw. Responses were collected between April 2023 and August 2023.

### Measures

#### Demographics

Participants provided basic demographics, including age, gender, ethnicity, and country of residence. They also self-reported whether they had ever been diagnosed with a mental health issue.

#### Psychometrics

The following psychometrics were measured to assess convergent validity: the Patient Health Questionnaire 9 (PHQ-9)[28] to measure depression, the Generalized Anxiety Disorder 7 (GAD-7)[29] to measure anxiety, the Perceived Stress Scale 4 (PSS-4)[30] to measure chronic stress, the Depression Anxiety and Stress Scale 21 (DASS-21)[31] to measure anxiety, depression, and stress in an integrated scale, and the Kessler Psychological Distress Scale (K-10)[32] to measure total levels of distress.

The Big Five Inventory (BFI) extraversion subscale[33] and the Emotion Regulation Questionnaire (ERQ)[34] were measured to assess divergent validity. Lastly, concurrent validity was assessed using the Patient Assessment of Upper Gastrointestinal Disorders-Quality of Life (PAGI-QOL)[35], which measures the quality of life in patients with upper gastrointestinal disorders. These scales were chosen as they are some of the most commonly used questionnaires to assess depression, anxiety, stress, personality, and quality of life in healthcare settings.

#### AGBW Survey feedback form

The optional feedback form asked participants to rate the AGBW Survey on a visual analogue scale for the following attributes; 1) how easy the questionnaire is to complete on a scale of 0(very hard) to 100(very easy), 2) how easy the questions are to understand on a scale of 0(very hard) to 100(very easy), and 3) how helpful they thought the scale would be for their gastric clinician to understand their mental wellbeing and provide them with more holistic care on a scale of 0(very unhelpful) to 100(very helpful). Participants were also asked to answer yes, no, or unsure to the question, “Would you like to see this questionnaire incorporated as part of routine assessment for your stomach symptoms, alongside medical testing?”

### Statistical Analysis

Details about the statistical analysis procedure and predefined acceptability criteria for the confirmatory factor analysis, reliability, and validity are detailed in the Supplementary Methods.

## Results

### Phase 1. Concept Selection

10 key concepts were identified as the most important indicators of mental health in this sample of patients with chronic gastroduodenal symptoms. These concepts were derived from specific items across three scales: items 1, 2, 4, and 7 of the PHQ-9; items 1, 2, and 3 of the PSS-4; and items 1, 2, and 7 of the GAD-7 (Figure 2). Further detail about how these 10 concepts were selected is provided in the Supplementary Results.

**Figure 2.**
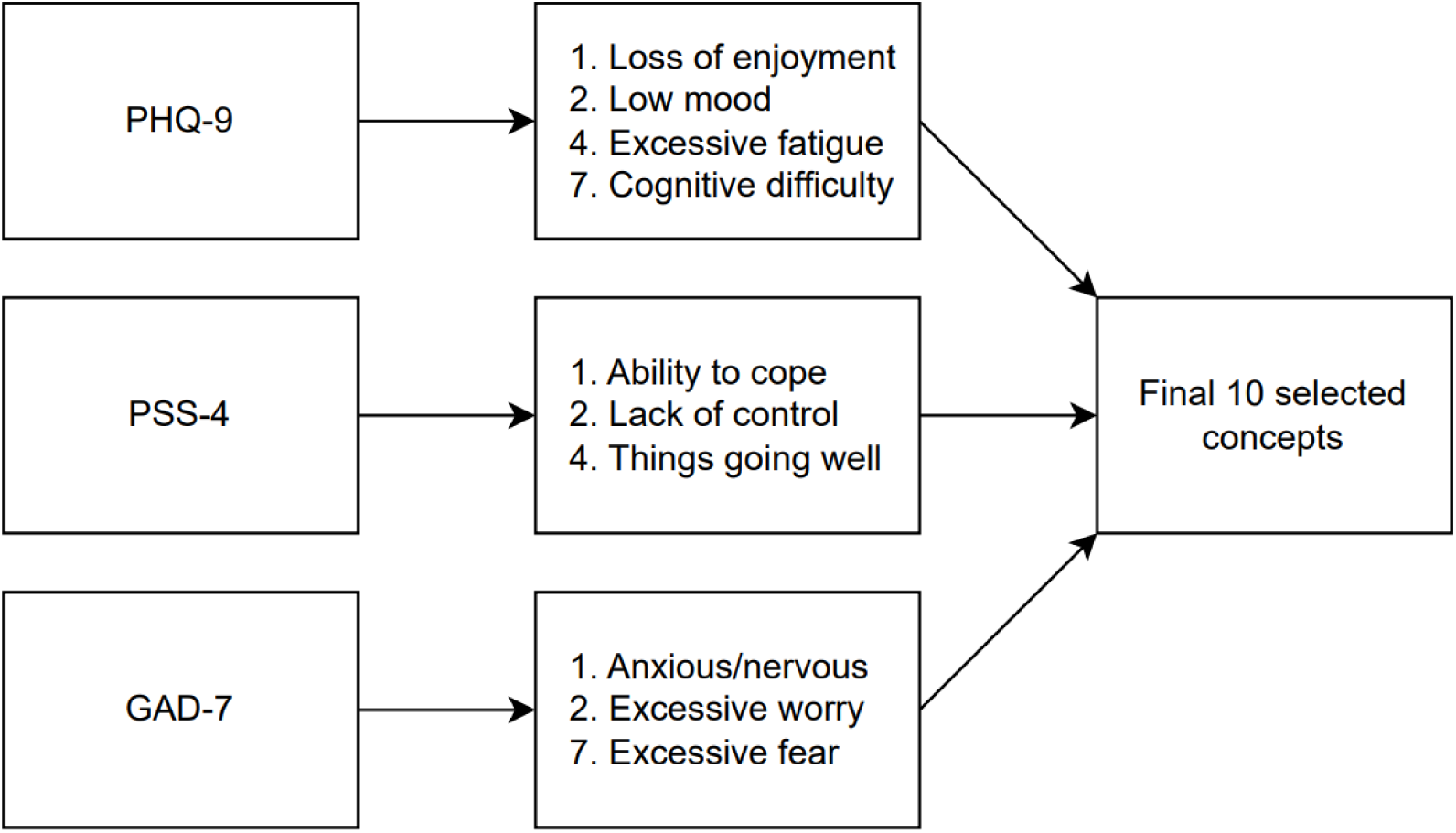
Flowchart showing the concepts selected from the original psychometrics.

### Phase 2. Item Generation

The expert panel drafted 10 closed-ended questions to cover the 10 concepts identified in Phase 1. A 5-point response format was chosen based on recommendations that reliability increases with more scale points for unipolar items, with diminishing returns after 5 points[36,37]. Similar to the PHQ-9 and GAD-7, a recall period of two weeks was chosen as this matches the criteria in the Diagnostic and Statistical Manual of Mental Health Disorders (5th Ed.; DSM-5)[38] and is recommended by the FDA due to increased recall bias beyond two weeks[39].

Both patients and clinicians in the precursory interview study[25] iterated the importance of prefacing patients before mental health assessments to explain why this information is being gathered and to reduce stigma and reassure them that this data will not be used to dismiss medical care. Based on these recommendations, the expert panel drafted a short preface explaining how this scale can help clinicians develop a more holistic understanding of the patient’s condition, enabling them to deliver a more tailored management plan. The preface also communicates how the scale is not diagnostic and cannot be used to attribute their gastrointestinal symptoms to their mental health.

Lastly, an opt-out option was included at the end of the preface to allow patients to decline the survey. The responses from the precursory interviews[25] determined this to be an essential addition for patients concerned about how their clinician might misuse or misinterpret their responses. A follow-up optional, open-ended question is provided for patients to leave a comment about why they have chosen not to answer this survey.

### Phase 3. Pre-Testing and Expert Feedback

Two rounds of feedback were gathered. Overall, the scale had high content and face validity and was seen as acceptable, easy to understand and complete, and useful to help clinicians further understand a patient’s condition and aid in developing tailored management. Both patients and clinicians were supportive of the use of the scale within clinical care.

Based on the feedback, the wording of the preface and scale items were edited to ensure clarity and ease of answering. The preface was expanded to include more information about the gut-brain axis to help patients further understand why they are being asked about their mental health. Lastly, an optional open-ended question was added to the end of the scale to allow patients to include additional comments regarding their mental wellbeing, enabling their clinician to develop a more comprehensive understanding of the survey results.

### The Final Scale: The Alimetry® Gut-Brain Wellbeing (AGBW) Survey

The final scale (see Supplementary File) consists of a patient preface, 10 closed-ended questions, and an optional 11th open-ended question. The patient preface explains to patients why these questions are being asked and how the data is being used. The 10 closed questions ask patients to rate how often they have felt or behaved in a certain way over the last two weeks on a scale of 0(none of the time) to 4(all of the time). Items 5 and 7 are written in a positive frame and must be reverse-coded. The scores from each question can be totalled to create a total gut-brain wellbeing score (out of 40). Three subscales can also be calculated; a depression subscale score made up of the total of questions 1-4 (out of 16), a stress subscale score made up of the total of questions 5-7, after reverse coding questions 5 and 7 (out of 12), and an anxiety subscale score made up of the total of questions 8-10 (out of 12). Higher scores indicate worse mental health. The scale concludes with an optional open-ended question asking patients to add any further comments about their mental wellbeing.

### Phase 4. Psychometric Validation

#### Sample Characteristics

A total of 311 participants completed the validation survey (mean age= 38.40 years, *SD*= 14.21, range= 18-76 years). Most respondents were female, white, and from the USA or British Commonwealth (Table 1). The Other countries included France (*n*= 1), Chile (*n*= 1), the Netherlands (*n*= 1), Ireland (*n*= 1), and Puerto Rico (*n*= 1). There was large overlap between the Rome diagnoses, with most patients meeting the Rome IV criteria for functional dyspepsia and chronic nausea and vomiting syndrome. Only 14% of respondents met the Rome IV criteria for only one gastroduodenal DGBI, with the majority (*n*= 186, 60%) meeting the criteria for three or more. Most patients had a self-reported previous diagnosis of anxiety or depression, with 74% self-reporting a previous diagnosis of any mental health disorder.

**Table 1.** Demographic characteristics of the survey respondents (*N*= 311)

|  | n (%) |
| --- | --- |
| Gender |  |
| Female | 262 (84%) |
| Male | 38 (12%) |
| Gender diverse | 11 (4%) |
| Country |  |
| United States | 120 (39%) |
| New Zealand | 89 (29%) |
| Australia | 43 (14%) |
| Canada | 29 (9%) |
| United Kingdom | 25 (8%) |
| Other | 5 (2%) |
| Ethnicity |  |
| White | 259 (83%) |
| Black | 5 (2%) |
| Asian | 13 (4%) |
| Hispanic | 14 (5%) |
| Māori/Pasifika | 12 (4%) |
| Other | 8 (3%) |
| Rome IV Criteria<sup>a</sup> |  |
| Functional dyspepsia | 260 (84%) |
| Chronic nausea and vomiting syndrome | 246 (79%) |
| Cyclic vomiting syndrome | 119 (38%) |
| Cannabinoid hyperemesis syndrome | 7 (2%) |
| Rumination syndrome | 115 (37%) |
| Belching disorder | 126 (41%) |
| Diagnosis of gastroparesis | 168 (54%) |
| Previous mental health diagnoses |  |
| Depression | 182 (59%) |
| Anxiety | 192 (62%) |
| Other | 62 (20%) |
| None | 82 (26%) |
| Prefer not to say | 4 (1%) |

Notes: n= number of respondents in that category; %= percentage of respondents in that category; <sup>a</sup> Rome IV criteria diagnosis is based on meeting the symptom criteria only; respondents can meet more than one Rome IV diagnosis.

### Descriptive Statistics

The 10 questions took participants an average of 87 seconds to complete (*IQR*= 58-107 seconds). As shown in Table 2, the full range of subscale scores available were used, except for the stress subscale, where no patient scored a 0/12. However, all individual questions used the whole range of answers (0-4). All three subscale scores and the total score were normally distributed, as shown by the lack of skew and kurtosis in Table 2 and the scale distributions in Figure 3. The mean and median scores of each subscale and the total score were in the middle of the scale range, further demonstrating normality.

**Figure 3.**
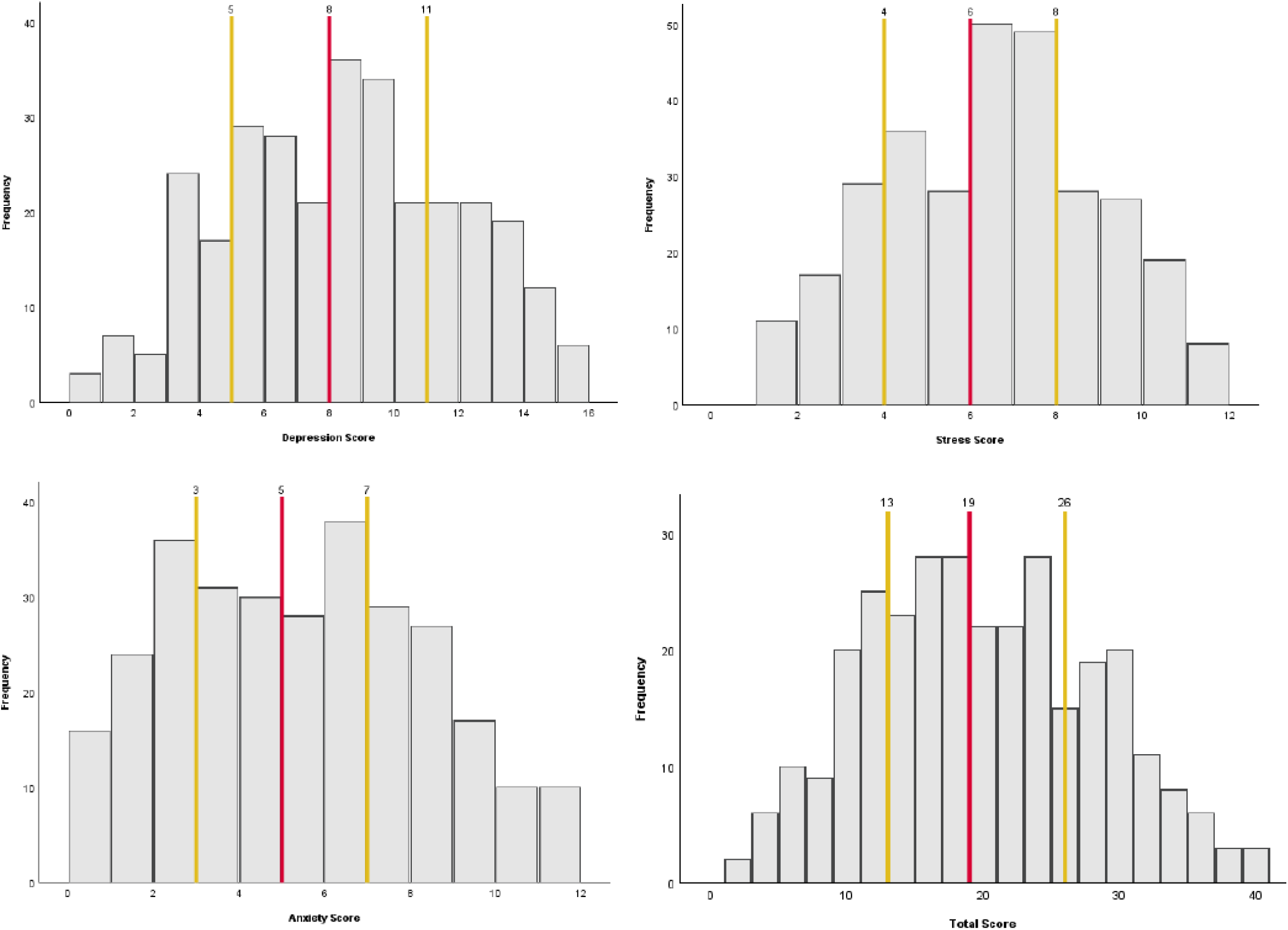
Histograms showing the distribution of the AGBW Survey subscale and total scores across the sample. The red line indicates the median score and the yellow lines indicate the interquartile ranges.

**Table 2.**
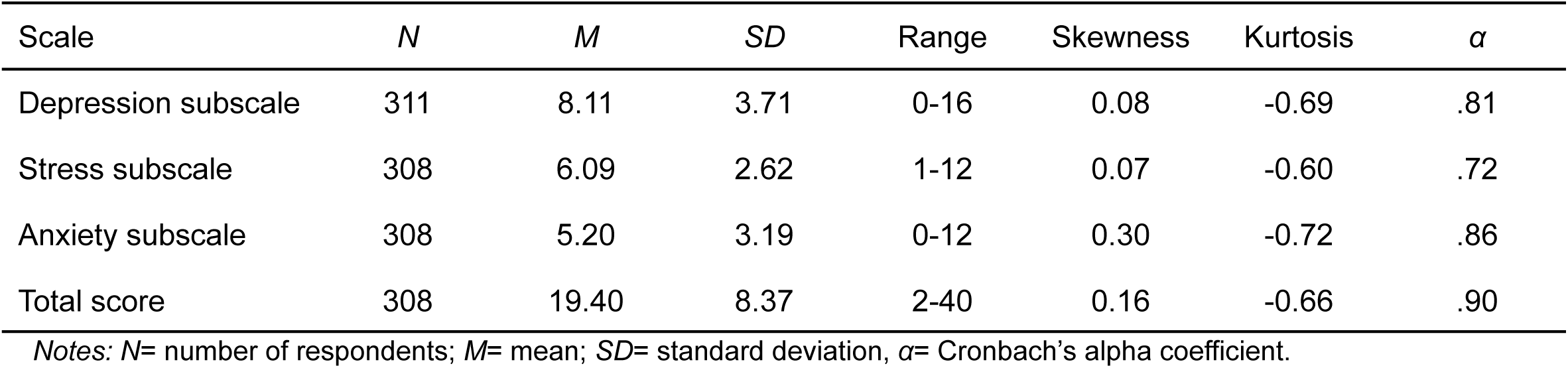
Descriptive statistics of the AGBW Survey’s subscales and total scores.

| Scale | *N* | *M* | *SD* | Range | Skewness | Kurtosis | $\alpha$ |
| --- | --- | --- | --- | --- | --- | --- | --- |
| Depression subscale | 311 | 8.11 | 3.71 | 0-16 | 0.08 | -0.69 | .81 |
| Stress subscale | 308 | 6.09 | 2.62 | 1-12 | 0.07 | -0.60 | .72 |
| Anxiety subscale | 308 | 5.20 | 3.19 | 0-12 | 0.30 | -0.72 | .86 |
| Total score | 308 | 19.40 | 8.37 | 2-40 | 0.16 | -0.66 | .90 |

Notes: *N*= number of respondents; *M*= mean; *SD*= standard deviation, $\alpha$ = Cronbach's alpha coefficient.

### Confirmatory Factor Analysis

The confirmatory factor analysis revealed that the three-factor model, splitting the depression, stress, and anxiety questions into separate factors, had a good fit, meeting all predefined acceptability criteria, as detailed in the Supplementary Methods (*χ^2^*_(32)_= 76.00, *p*<.001, *χ^2^/df*= 2.37, NFI= .95, CFI= .97, TLI= .96, SRMR= .04, RMSEA= .07 [.05-.09]).

### Reliability

As shown in Table 2, the Cronbach’s *α* coefficients demonstrated excellent internal consistency reliability for the AGBW Survey total score, good internal consistency reliability for the anxiety and depression subscale scores, and acceptable internal consistency reliability for the stress subscale score. In the total score and all subscales, eliminating additional items resulted in no substantial increases in reliability. The inter-item correlations ranged from *r*= .31-.75, indicating good consistency between the individual items. The corrected item-total correlations ranged from *r*= .53-.78, indicating good consistency between the individual items and the subscale and total scores.

### Validity

#### Convergent validity

Most of the correlations between the AGBW Survey’s subscale scores and total scores and the scales used to assess convergent validity showed significant correlations with large effect sizes over *r*= .50 (Table 3), indicating good convergent validity. However, the correlation between the AGBW stress subscale and the DASS-21 stress subscale was just under *r*= .50, indicating acceptable convergent validity despite the high correlation with the PSS-4.

**Table 3.** Pearson correlation coefficients between the AGBW Survey scores and comparative questionnaires used for validity testing.

|  |  | Depression subscale | Stress subscale | Anxiety subscale | Total score |
| --- | --- | --- | --- | --- | --- |
| Convergent validity | PHQ-9 total | .77* |  |  |  |
|  | PSS-4 total |  | .80* |  |  |
|  | GAD-7 total |  |  | .78* |  |
|  | DASS-21 depression subscale | .70* |  |  |  |
|  | DASS-21 stress subscale |  | .46* |  |  |
|  | DASS-21 anxiety subscale |  |  | .57* |  |
|  | K-10 total |  |  |  | .87* |
| Divergent validity | BFI extraversion subscale | -.26* | -.27* | -.20* | -.28* |
|  | ERQ cognitive reappraisal subscale | -.32* | -.32* | -.24* | -.34* |
|  | ERQ expressive suppression subscale | .22* | .19* | .20* | .23* |
| Concurrent validity | PAGI-QoL total | .64* | .52* | .55* | .66* |
|  | PAGI-QoL psychological wellbeing and distress subscale | .74* | .64* | .68* | .79* |

Notes: PHQ-9= Patient Health Questionnaire 9; PSS-4= Perceived Stress Scale 4; GAD-7= Generalized Anxiety Disorder 7; DASS-21= Depression Anxiety and Stress Scale 21, K-10= Kessler Psychological Distress Scale; BFI= Big Five Inventory, ERQ= Emotion Regulation Questionnaire; PAGI-QoL= Patient Assessment of Upper Gastrointestinal Disorders-Quality of Life; \* denotes significance at $p < .05$ .

#### Divergent validity

The correlations between the AGBW subscale and total scores and the BFI and ERQ scores all demonstrated small effect sizes, which were considerably lower than the convergent validity correlations (Table 3), demonstrating successful divergent validity.

#### Concurrent validity

All correlations between the AGBW subscale and total scores and the PAGI-QoL total scores and psychological wellbeing and distress subscale scores were statistically significant and over *r*= .50, demonstrating large effect sizes and therefore providing strong evidence of concurrent validity (Table 3).

#### Known groups validity

As shown in Table 4, there was a significant difference between those with and without any previous mental health diagnosis, with very large effect sizes. Those with a previous mental health diagnosis scored on average higher on all three subscale scores and the AGBW Survey total score than those who did not have a previous mental health diagnosis, indicating good known groups validity.

**Table 4.** Results from the independent samples t-tests used for known groups validity of the AGBW Survey.

|  |  | Depression subscale | Stress subscale | Anxiety subscale | Total score |
| --- | --- | --- | --- | --- | --- |
| Previous mental health diagnosis | Yes, M (SD) | 9.18 (3.39) | 6.74 (2.40) | 6.07 (3.03) | 22.00 (7.50) |
|  | No, M (SD) | 5.38 (2.97) | 4.44 (2.39) | 2.99 (2.37) | 12.71 (6.40) |
|  | *p*-value | <.001* | <.001* | <.001* | <.001* |
|  | Cohen's *d* | 1.16 | 0.96 | 1.07 | 1.28 |
| Gender | Female, M (SD) | 7.96 (3.65) | 6.06 (2.61) | 5.16 (3.26) | 19.18 (8.36) |
|  | Male, M (SD) | 8.42 (3.96) | 6.14 (2.81) | 5.03 (2.69) | 19.54 (8.58) |
|  | *p*-value | .237 | .437 | .405 | .405 |
|  | Cohen's *d* | 0.12 | 0.03 | 0.04 | 0.04 |

Notes: M=mean; SD=standard deviation; *p*-values were calculated using independent samples t-tests; \* denotes significance at $p < .05$ .

Additionally, independent samples t-tests showed that those patients who had a previous depression diagnosis had significantly higher scores on the AGBW depression subscale (*M*= 9.44, *SD*= 3.46), than those who did not have a previous depression diagnosis (*M*= 6.31, *SD*= 3.19, *p*< .001, *d*= 0.93), with a large effect size. Similarly, those who had a previous diagnosis of an anxiety disorder had higher scores on the AGBW anxiety subscale (*M*= 6.32, *SD*= 3.03) than those who did not have a previous anxiety diagnosis (*M*= 3.45, *SD*= 2.52, *p*< .001, *d*= 1.01), with a large effect size.

In contrast, there were no significant differences between males and females for any of the subscale or total scores (Table 4), as was hypothesised in the predefined acceptability criteria[40–43].

### Feedback Form

240 patients completed the optional feedback form. The scale questions were rated as easy to complete and easy to understand, with an average score of 85.13 (*SD*= 18.48) and 85.77 (*SD*= 17.51) out of 100, respectively. The patients also rated the questions as helpful for their gastric clinician to understand their mental wellbeing and provide them with more holistic care, with an average score of 70.24 (*SD*= 26.36) out of 100. Lastly, although 33% were unsure if they would like to see these questions incorporated as part of the routine assessment for their stomach symptoms, only 10% answered no, and 57% indicated yes, suggesting the overall acceptability of these questions.

## Discussion

This research used a multi-phase, mixed-methods process, incorporating co-design with patients and clinicians to develop and validate a brief, novel mental health scale for patients with chronic gastroduodenal symptoms.

The AGBW Survey was developed to be a novel addition to the existing pool of psychometrics for patients with chronic gastroduodenal symptoms, complementing other mental wellbeing tools that measure gastrointestinal symptom-specific anxiety[44] and quality of life[35]. Due to the high presence of psychological comorbidities[13,19] and clinical recommendations for routine psychological assessments in these patients[9,13,45], clinicians have typically relied on general mental health scales. However, these assessments are not contextualized for use within patients with chronic gastroduodenal symptoms and may potentially exaggerate the reporting of mental health issues due to the wording of items that emphasize physical symptomatology[13]. In contrast, the AGBW Survey is designed to focus more specifically on mental health concepts relevant to patients with chronic gastroduodenal symptoms.

Furthermore, the AGBW Survey is multidimensional and combines assessments of depression, anxiety, and stress into a single, brief scale, with confirmatory factor analysis supporting the presence of this three-factor model. This scale allows a quick assessment of a patient’s general wellbeing as well as more specific dimensions of common mental health issues. This scale’s brief nature reduces the clinician and patient burden associated with psychological assessment batteries, which is particularly important as gastroenterology clinicians often report time constraints as the key reason for not regularly assessing a patient’s mental wellbeing[25]. The AGBW Survey is intended to complement routine medical tests, such as body surface gastric mapping[46,47], to provide more integrated evaluations and management plans. Research has demonstrated that incorporating psychological support for patients with chronic gastroduodenal symptoms as part of multidisciplinary care leads to better care and more effective symptom management[48–51]. Therefore, the results from the AGBW Survey could prompt clinicians to consider incorporating psychological referrals or interventions into the patient’s care plan alongside traditional medical care.

The AGBW Survey demonstrated excellent reliability and good validity, including strong correlations with existing mental health measures, indicating its suitability for clinical use. The scale also successfully discriminated between patients with and without a self-reported mental health diagnosis. Similar results were also seen for the three individual subscale scores. However, the stress subscale did show lower, but still acceptable, internal consistency reliability and convergent validity. This finding is likely related to the fact that the stress subscale included two reverse-coded items, which, by nature, can lead to lower reliability[52,53]. Additionally, the DASS-21 stress subscale was only moderately correlated with both the AGBW stress subscale and the PSS-4, suggesting that the lower convergent validity may be attributable to the DASS-21, rather than the AGBW subscale.

The scale’s psychometric properties demonstrated a normal distribution with little to no skew. Generally, psychometrics are positively skewed, with most respondents scoring on the lower end[54,55]. However, patients with chronic gastroduodenal symptoms often experience higher psychological comorbidities than the general population[10,11,56,57], which may account for the high scores and lack of skew. This finding was further emphasized by the high number of participants with a previous mental health diagnosis, which further highlights the importance of routinely assessing mental wellbeing in this patient population.

Contrary to our hypothesis, the total and subscale scores were unable to discriminate between males and females, which could be attributed to the low number of males recruited. Males are less likely than females to have a functional gastrointestinal disorder[1] and participate in online survey-based research[58,59], so the limited male representation is unsurprising, even despite researcher efforts to boost male participation. Evidence suggests that males with functional gastrointestinal disorders are less depressed, anxious, and stressed than females, regardless of their symptom severity[43,60]. However, in the current study, males had relatively high scores on the scale, potentially due to self-selection bias. Despite this, the pre-defined acceptability criteria (detailed in the Supplementary Methods), which were based on existing guidelines[61,62], detailed that at least or equal to 75% of the validity hypotheses must be met to demonstrate good construct and criterion validity. Even with the lack of successful gender discrimination, 90% of the validity hypotheses were met, indicating successful validation.

A novel aspect of this scale is the addition of the patient preface, opt-out option, and final open-ended question. These were important additions suggested by clinicians and patients during the precursory interviews[25] and feedback phase to increase the scale’s acceptability and usability, and decrease stigma. In particular, the open-ended question allows patients to provide valuable context to their survey responses, providing clinicians with important additional information that can be used for targeted clinical management plans and reducing the chance of the patient’s symptoms being dismissed as exclusively psychological.

Strengths of this scale development process include the co-development with patients, clinicians, and interdisciplinary experts throughout every phase, which ensured the development of an acceptable, understandable, useful, and clinically relevant scale. Furthermore, due to the international approach, the samples included patients from diverse countries and health contexts. However, most patients were from Western countries and self-identified as ethnically white, which may restrict the generalisability of these results. Previous research has shown the invariance of existing mental health scales across ethnicities, such as the PHQ-9[63,64] and GAD-7[65]. Therefore, it is expected the AGBW Survey should have a similar ethnic invariance. With the inclusion of 7% US ethnic minorities, we expect the results to be generalisable in the US, although additional ethnicity contexts were not addressed. Furthermore, the scale is currently only available in English and still needs to be translated and validated in other languages.

Additionally, as the validation survey was cross-sectional, we could not assess the scale’s predictive validity. Research is currently underway to evaluate whether the combination of the results from the AGBW Survey and physiological tests, such as body surface gastric mapping[46,47], can help predict which patients benefit from integrated care. This research will further inform the scale’s clinical utility as an aid to guide case formulation and clinical management decisions.

### Summary

The AGBW Survey was developed using a multi-phase, co-design process that included precursory interviews, consensus from an interdisciplinary panel of experts, and feedback from clinicians, patients, and key opinion leaders. This research demonstrates that the AGBW Survey is a well-accepted, valid, and reliable scale for assessing mental health in patients with chronic gastroduodenal symptoms. This scale combines assessments of depression, stress, and anxiety, using items contextualized for patients with chronic gastroduodenal symptoms, allowing a brief assessment of a patient’s mental health. While not designed for diagnostic purposes, the AGBW Survey can be incorporated into routine clinical testing to complement existing physiological tests to provide more integrated evaluations and management plans. Moreover, its utility extends to research contexts, where it can be used to assess a patient’s mental health at baseline or evaluate changes over time.

## Conflicts of interest

GOG and AG hold grants and intellectual property in the field of gastrointestinal electrophysiology and are Directors in Alimetry Ltd. GOG is also a Director in The Insides Company. ML, IP, GS, GS, PD, CD, CNA, and SC are members of Alimetry Ltd. GH holds options in Alimetry. The remaining authors have no relevant conflicts to declare.

## Funding

This study is funded by a New Zealand Health Research Council Programme Grant 3715588.

## Data Availability

The data that support the findings of this study are available from the corresponding author, upon reasonable request.

## Acknowledgements

This research was conducted on behalf of the BSGM consortium. We thank India Fitt and Gen Johnson for their invaluable research assistance.

## Supplementary Methods

### Phase 1. Concept Selection

The precursory interview study[25] identified depression, stress, and anxiety as the most important mental health domains to assess clinically in the target patient group. Both patients and clinicians desired a brief scale that combined these three domains. Based on this feedback, validated concepts from existing and widely used depression, stress, and anxiety questionnaires were used to identify the most important concepts for patients with chronic gastroduodenal symptoms. Using existing questionnaires as the basis for concept identification ensured content validity in the new questionnaire.

### Design

Concept selection involved a combination of expert feedback and the analysis of psychometric data gathered from a sample of patients with chronic gastroduodenal symptoms, which was collected as part of another multi-national consortium database study, on behalf of the BSGM working group (https://www.bsmconsortium.com/). Data collection was conducted in Auckland (New Zealand), Calgary (Canada), and Western Sydney (Australia). Ethics were obtained for each data collection site via: The Auckland Health Research Ethics Committee (AHREC; AH1130), The University of Calgary Conjoint Health Research Ethics Board (REB19-1925), and the Human Research Ethics Committee at Western Sydney (H13541). All participants provided written informed consent.

### Sample

The sample consisted of 79 patients (73% female; mean age= 38.1 years, age range= 15-81 years) and 50 healthy controls (62% female; mean age= 39.2 years, age range= 19-84 years). Patients were defined as meeting the Rome IV criteria[27] for functional dyspepsia and/or chronic nausea and vomiting syndrome, whilst healthy controls were defined as having no chronic gastroduodenal symptoms.

### Procedure

Participants completed a battery of psychometric questionnaires while participating in another study, using methods described elsewhere[47]. The Patient Health Questionnaire 9 (PHQ-9)[28] was used to measure levels of depression, the Perceived Stress Scale 4 (PSS-4)[30] was used to measure levels of stress, and the Generalised Anxiety Disorder 7 (GAD-7)[29] was used to measure levels of anxiety. These questionnaires were included as they are some of the most widely used and well-validated existing mental health assessment tools and are frequently used in patients with chronic gastroduodenal symptoms[66,67].

### Statistical Analysis

The key concepts from each questionnaire were selected by statistically evaluating the weighting of the concept as a total predictive value to the original scales’ total score, using the following methodology in Python v3.7.

Firstly, the sample of 79 patients was separated into two subsamples: a 75% training sample and a 25% validation sample. Using bootstrapping 1000 random samples with replacement were generated from the original 75% cohort. The 1000 samples were then passed through a univariate feature selection to rank the importance of each item. Iteratively working from one feature to all features, the combination of features was used to train a regression model and then the predicted scores were compared to the actual scores. Results were compiled such that 1000 predictive scores were amassed for each feature value, along with the mean and 95% confidence interval. The subscale of each scale was then determined by allowing in the number of features required to have a mean accuracy of 90%. In the event of edge cases being close to the threshold, feature weighting was compared statistically (using a two-tailed t-test) to ensure each scale weighting was statistically different (no correction for multiple comparisons was performed). The defined subset of items was then trained on the original 75% sample and validated against the testing 25% sample. A valid score was reported if the R^2^ value was greater than 0.9.

An additional confirmatory analysis was conducted to confirm the specificity of the extracted questions to patients compared to healthy controls. This analysis involved repeating the procedure described above in the healthy control group (n= 50). The rankings of the questions were compared between the two groups using two-tailed t-tests.

### Phase 4. Psychometric Validation

#### Statistical Analysis

Data were analyzed using IBM SPSS Statistics v29. A *p*-value of .05 was considered statistically significant. Partial responses were included in the study as long as the patient had completed at least the demographics questions and the first four questions (the depression subscale) of the AGBW Survey. As a result, the validity and reliability calculations vary in terms of the number of respondents included, with a minimum of *N*= 295 within each calculation.

##### Confirmatory factor analysis

Confirmatory factor analysis with the maximum likelihood estimation method was conducted using IBM SPSS AMOS v26 using a three-factor model, splitting the depression, stress, and anxiety questions into separate factors. The model’s goodness of fit was evaluated using multiple indices: chi-square/degree of freedom (*χ^2^/df*), the normed fit index (NFI), the comparative fit index (CFI), the Tucker Lewis Index (TLI), the standardized root mean square residual (SRMR), and the root mean square error of approximation (RMSEA). An acceptable model of fit was predefined as a *χ2/df*< 5, an NFI> .95, a CFI> .90, and TLI> .95, an SRMR< .08, and an RMSEA<.08[68–70].

##### Reliability

Cronbach’s alpha coefficients (α) were calculated to examine the internal consistency reliability of the subscale and total scores[71]. A value of α> .70 indicates acceptable reliability, α> .80 ideal reliability, and α> .90 excellent reliability[36,61,71,72]. Inter-item correlations and corrected item-total correlations were calculated to assess the correlations between the scale items and the correlations between each item and the subscale scores/total score without that item, respectively. A value of *r*> .30 indicates good consistency between the scale items (for inter-item correlations) and the subscale/total scores (for item-total correlations)[36,71].

##### Validity

To demonstrate good construct and criterion validity for the subscale and total scores, at least or equal to 75% of the hypotheses below for convergent, divergent, concurrent, and known-groups validity were required to be met for each subscale/total score[61,62].

The convergent validity of the AGBW depression subscale was assessed using Pearson’s correlation coefficients with the PHQ-9 total score and the depression subscale score of the DASS-21; the AGBW stress subscale was compared with the PSS-4 total score and the DASS-21 stress subscale score; the AGBW anxiety subscale was compared against the GAD-7 total score and the DASS-21 anxiety subscale score; and lastly, the AGBW total score was compared against the K-10 total score. A value of *r*> .50 indicates good convergent validity and *r*> .30 acceptable convergent validity[62,71,73].

Divergent validity was analyzed using Pearson’s correlation coefficients between the AGBW subscale and total scores, and the BFI extraversion subscale score, the ERQ cognitive reappraisal subscale score, and the ERQ expressive suppression subscale score. Divergent validity was achieved if these correlation coefficients were weaker than the correlations with the scales used for convergent validity[71,74].

Concurrent validity was analyzed using Pearson’s correlation coefficients between the AGBW subscale and total scores and the PAGI-QoL total score and psychological wellbeing and distress subscale score. Concurrent validity was achieved if these correlation coefficients were positive and statistically significant; with r> .50 indicating strong, r= .30-.50 moderate, and r< .30 weak evidence.

Known groups validity was analyzed using one-tailed independent samples t-tests between groups who theoretically should have different scores on the scale[36]. Patients with a previous mental health diagnosis were expected to have significantly higher average scores on the AGBW subscale and total scores than those without a previous mental health diagnosis. Females were also expected to have significantly higher average subscale and total scores than males[40–43].

## Supplementary Results

### Phase 1. Concept Selection

During the evaluation of the PHQ-9, results showed five potential factors (items 1, 2, 4, 7, 8) were required to meet the desired threshold of 0.9. However, there was only a marginal difference in accuracy between including either four or five of these items, with accuracy values shifting from 0.89 to 0.9. Upon applying statistical criteria, a t-test was conducted comparing the weighting scores of these items, with a significant difference revealed. Coupled with expert recommendations suggesting the exclusion of questions related to physical symptomatology for this patient population, item 5 (poor appetite/overeating) was ultimately excluded from the scale.

The results from the confirmatory analysis with healthy controls indicated that all scale values were significantly different, providing evidence to support the assertion that this scale is unique and contextualised to patients with chronic gastroduodenal symptoms. This selection of these items increased the specificity and contextualisation of the questions for use in patients with chronic gastroduodenal symptoms, while also allowing for a reduction in clinician and patient burden by including a smaller subset of questions than would typically be completed in separate anxiety, stress, and depression questionnaires.

## Alimetry® Gut-Brain Wellbeing (AGBW) Survey

The following 10 questions will ask you about your mental wellbeing over the past 2 weeks.

Research has shown that stomach symptoms and mental wellbeing can affect each other through the gut-brain axis, a physical connection between the stomach and brain.

These questions will help your clinician better understand your mental wellbeing in connection with your symptoms and stomach’s activity. This allows for a more holistic understanding of your condition, enabling a more personalised management plan.

Please note these questions cannot be used to diagnose you with a mental health condition and are not intended to attribute your symptoms to your mental health. Your answers are confidential and will be seen by your referring clinician in your test report.

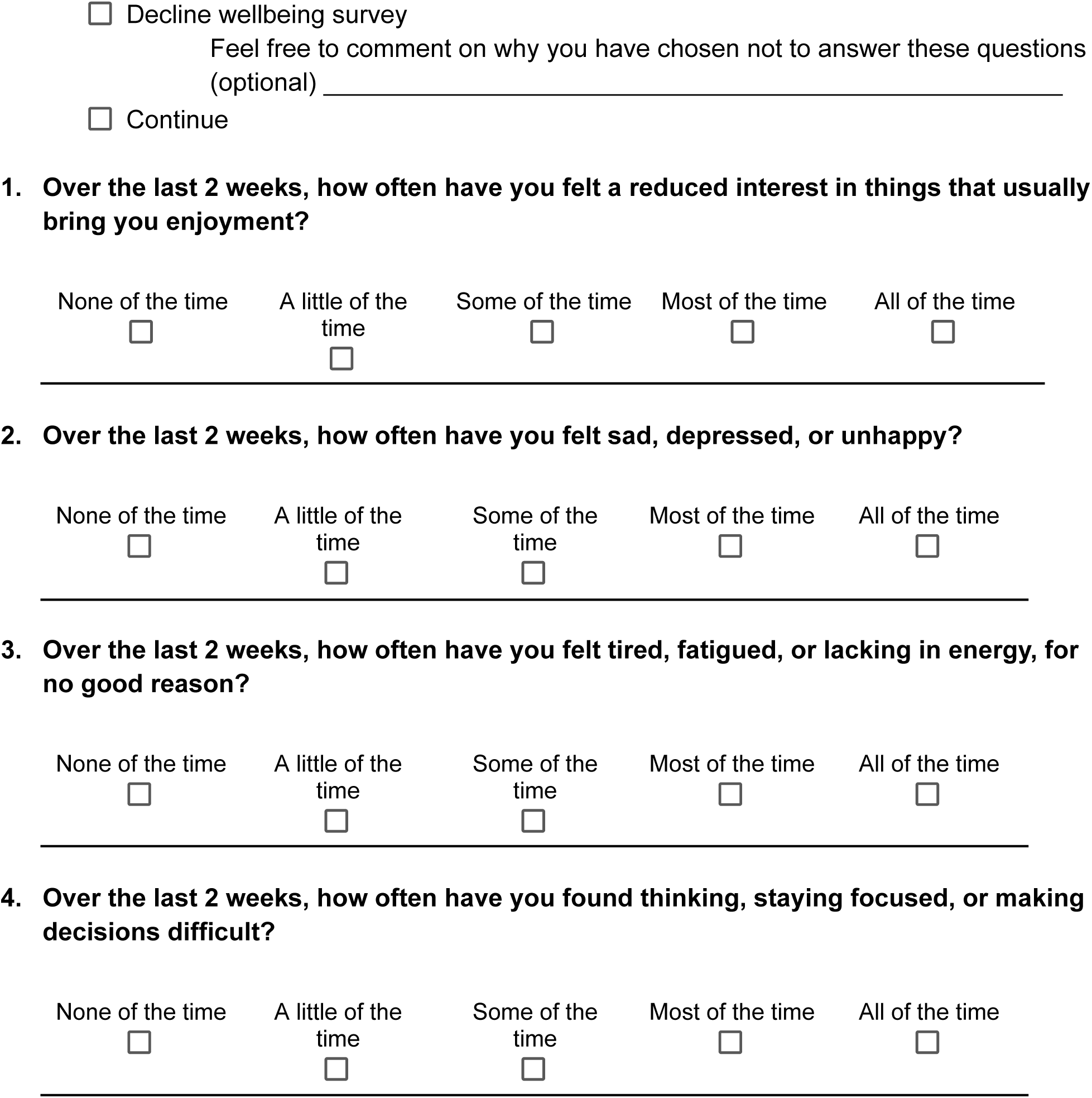
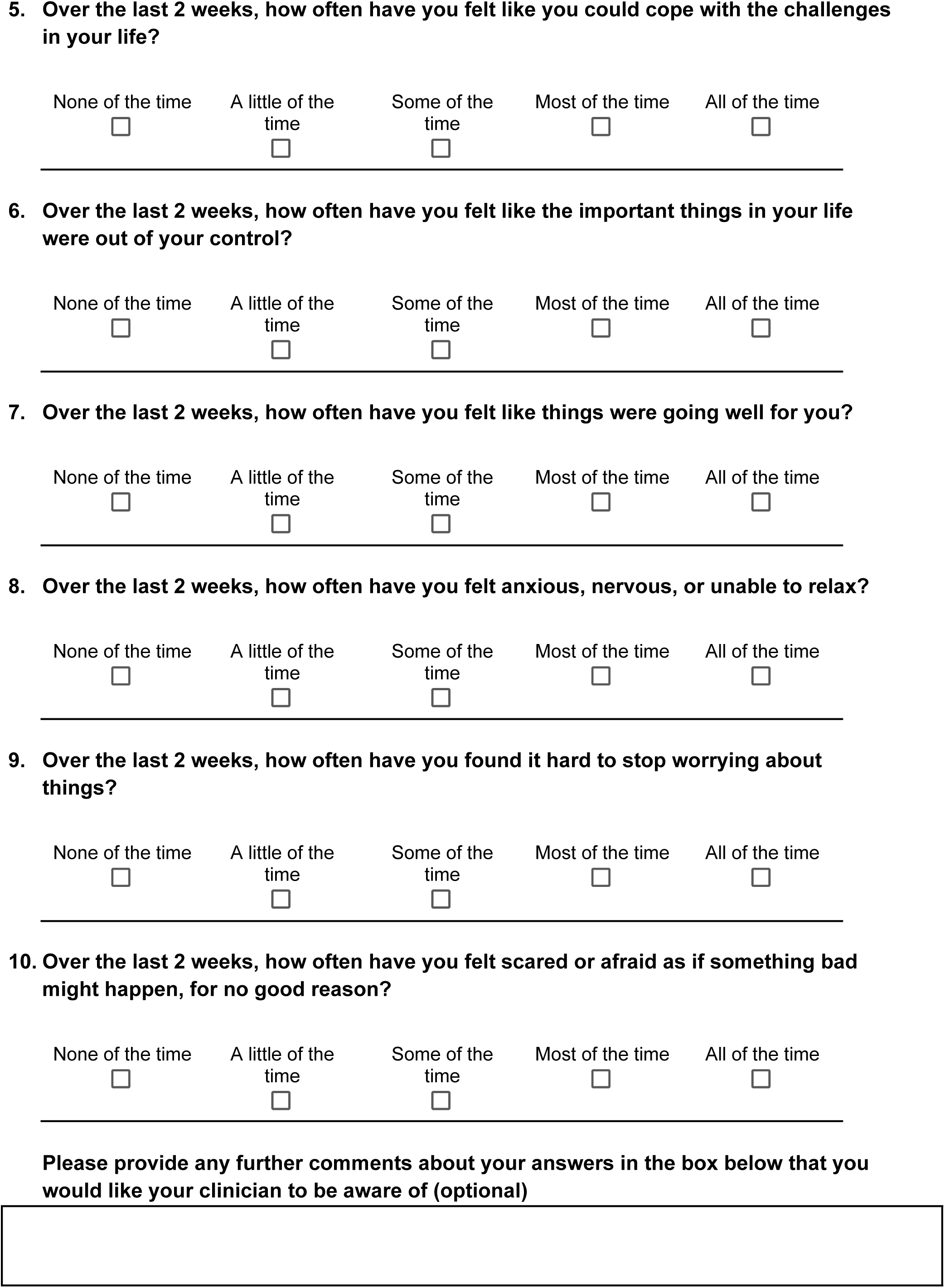

## Notes

### Clinical Trial

ACTRN12623000385640

### Author Declarations

Ethics was obtained via the Auckland Health Research Ethics Committee (AHREC; AH1130), The University of Calgary Conjoint Health Research Ethics Board (REB19-1925), and the Human Research Ethics Committee at Western Sydney (H13541).

### Summary of Updates

The manuscript has been revised to add in the validation of the scale alongside the development process. The manuscript has also been made more concise to fit the validation study.

